# Disaggregating the Genetic Overlap Between Educational Attainment and Substance Use Phenotypes

**DOI:** 10.1101/2025.07.18.25331735

**Authors:** Christal N. Davis, Yousef Khan, Zachary Piserchia, Joshua C. Gray, Henry R. Kranzler

## Abstract

**Background:** Educational attainment (EA) is positively genetically correlated with alcohol and cannabis use, but negatively correlated with alcohol and cannabis use disorders. These opposing associations suggest that shared genetic influences differ by level of substance involvement and for the cognitive (CogEA) and non-cognitive (NonCogEA) components of EA.

**Methods:** To test this, we examined the shared genetic architecture of EA, CogEA, and NonCogEA with alcohol consumption (AC), alcohol use disorder (AUD), lifetime cannabis use (CanUse), and cannabis use disorder (CUD). We used bivariate causal mixture models, local genetic correlation analyses, and conditional/conjunctional false discovery rate analyses to identify polygenic, regional, and variant-level overlap.

**Results:** For variants shared with AC, 48.12% and 52.86% showed concordant effects with CogEA and NonCogEA, respectively, compared to 38.40% and 41.02% for AUD. For CanUse, 71.42% and 65.56% of shared variants were concordant with CogEA and NonCogEA, respectively, versus 37.97% and 42.23% for CUD. Jointly associated loci were identified for each substance-EA pair. AUD and CogEA variants were enriched for expression in the basal ganglia, CUD and CogEA in the substantia nigra, and CanUse and NonCogEA in the cerebellum and its hemispheres.

**Conclusions:** Although EA is associated with greater substance use and lower risk for disorder, this pattern reflects a complex mix of concordant and discordant variant effects. CogEA and NonCogEA show partially distinct patterns, particularly for cannabis-related traits, highlighting the importance of disaggregating EA to clarify the genetic architecture underlying its paradoxical associations with substance-related traits.

## Introduction

Educational attainment (EA) is a complex, multifaceted trait influenced by cognitive abilities (e.g., intelligence) and non-cognitive attributes (e.g., perseverance and personality) (Demange et al., 2021). In many countries, including the United States, EA significantly influences socioeconomic status and physical and mental health (Demange et al., 2024; Kondirolli & Sunder, 2022; LaVeist et al., 2023). Given these far-reaching implications, a clearer understanding of how the genetic architecture of EA intersects with that of other complex traits, including substance use and substance use disorders (SUDs), could provide insight into the biological and behavioral pathways that link education to health.

Genetic associations between EA and substance use behaviors are at times paradoxical. Although EA is positively genetically correlated with alcohol and cannabis use (Kranzler et al., 2019; Pasman et al., 2018), it has an equally negative genetic correlation (*r*_g_) with alcohol use disorder (AUD) and cannabis use disorder (CUD) (Kranzler et al., 2019; Levey et al., 2023). One explanation for these findings is that genetic influences on EA promote experimentation or regular low-level substance use without increasing SUD risk. For example, genetic variants associated with openness and curiosity—both important for academic success (Chen et al., 2025)—may encourage experimentation with substances (Moncel et al., 2025), while other etiologic factors shape the transition to disorder. Similarly, higher socioeconomic status may increase individuals’ access to and opportunities for substance use while providing them with social and material resources that can buffer against the harmful consequences of use (Probst et al., 2020). A more complete understanding of these genetic associations requires moving beyond broad *r*_g_ to examine the contributions of specific genomic regions and individual variants to the patterns observed.

To this end, we conducted analyses of increasing genetic specificity to dissect the overlap between EA and four substance-related traits: alcohol consumption (AC) (Kember et al., 2023), lifetime cannabis use (CanUse) (Pasman et al., 2018), AUD (Kember et al., 2023), and CUD (Levey et al., 2023). First, we assessed the overall polygenic overlap for each trait pair— EA/AC, EA/CanUse, EA/AUD, and EA/CUD—using bivariate causal mixture models, which estimate the proportion of shared causal variants irrespective of their direction of effect (Frei et al., 2019; Holland et al., 2020). Next, we conducted Local Analysis of [co]Variant Associations (LAVA) (Werme et al., 2022) to identify genomic regions where EA shows significant *r*_g_ with each substance-related trait. We then performed conditional and conjunctional false discovery rate (cond/conjFDR) analyses (Andreassen et al., 2013) to identify individual genetic variants that contribute to the overlap between EA and substance use behaviors. Comparing the findings across traits and methods enabled us to identify patterns that differentiate the association of EA with substance use from its association with SUDs.

To refine our understanding, we performed similar analyses focusing on the cognitive (CogEA) and non-cognitive (NonCogEA) components of EA (Demange et al., 2021). Although a published GWAS showed no significant differences between CogEA and NonCogEA in their *r*_g_ with alcohol dependence, alcohol use, and cannabis use (Demange et al., 2021), we used methods that reveal subtle associations that may not be evident with *r*_g_. Collectively, this approach reveals the shared and distinct genetic pathways that link EA with substance use behaviors and enhances our understanding of the role of educational, cognitive, and noncognitive factors in shaping health behaviors among European-like genetic ancestry (EUR) individuals.

## Methods

### Summary Statistics

We conducted secondary analyses of summary statistics from large GWAS of EUR individuals for AC (N = 296,989) (Kember et al., 2023), AUD (N = 296,989, N_cases_ = 53,458) (Kember et al., 2023), CanUse (N= 162,082) (Pasman et al., 2018), and CUD (N = 445,847, N_cases_ = 22,260) (Levey et al., 2023; Pasman et al., 2018). The original studies received ethical approval and obtained informed consent from all participants.

In the Million Veteran Program, AC was assessed using the Alcohol Use Disorders Identification Test-Consumption (AUDIT-C) score (Bush et al., 1998), while AUD and CUD diagnoses were based on International Classification of Diseases 9^th^ Revision (ICD-9) and 10^th^ Revision (ICD-10) codes. The CanUse GWAS (Pasman et al., 2018) comprised samples from the UK Biobank and International Cannabis Consortium, with each cohort providing self-reported lifetime cannabis use information. Summary statistics were obtained from the largest GWAS of EA (Okbay et al., 2022), excluding the UK Biobank and 23andMe to avoid sample overlap (N = 324,162). We also obtained summary statistics from a GWAS-by-subtraction of EA and cognitive performance (CogEA N = 257,700 and NonCogEA N = 510,795) (Demange et al., 2021).

### Polygenic Overlap

We used MiXeR to estimate the overall polygenic overlap between substance-related traits and EA (Frei et al., 2019). We first used univariate models to estimate the polygenicity and discoverability of each trait (Holland et al., 2020). We ensured that the AIC values of the univariate models were positive, which indicates that there is adequate power for bivariate MiXeR. Bivariate models estimate the genetic overlap between traits, regardless of the direction of effect of causal variants. The Dice coefficient estimates the proportion of the total number of causal SNPs that are shared.

### Local *r*_g_

We used LAVA (Werme et al., 2022) to estimate the local *r*_g_ across 2,495 loci—each consisting of approximately 1-Mb blocks of the genome—with the 1000 Genomes Phase 3 EUR panel as a linkage disequilibrium (LD) reference (Auton et al., 2015). We used univariate association signals to estimate local SNP heritability and bivariate association signals to identify regions with significant local *r*_g_. For the univariate test, we applied a Bonferroni correction to account for multiple testing (*p* < 0.05/2,495), whereas for the bivariate test, we applied a false discovery rate (FDR) p-value correction.

### Joint SNP-Level Associations

To detect SNPs jointly associated with each pairwise combination of substance- and education-related traits, we conducted cond/conjFDR analyses (Smeland et al., 2020). First, we obtained conditional FDR (condFDR) estimates by conditioning the effect for one trait (e.g., AC) on the other trait (EA). This analysis re-ranks the test statistic for a primary trait based on the strength of association with a secondary trait. For each pair of traits, we performed condFDR twice by switching the primary and secondary traits to determine a conjFDR value, defined as the maximum of two condFDR values. This provides a conservative estimate of each SNP’s association with both traits. Test statistics were corrected using a genomic inflation control procedure that randomly prunes SNPs across 500 iterations using an LD r^2^ threshold of 0.1 (Smeland et al., 2020).

### SNP Annotation

We performed annotation of independent loci and identified functional SNP effects using Functional Mapping and Annotation (FUMA) v1.7.0 (Watanabe et al., 2017). SNPs with an LD block distance of ≤250 kb were merged. Independent significant SNPs were those with r^2^ < 0.6 and conjFDR < 0.05, whereas lead SNPs were those with r^2^ < 0.1. Candidate SNPs comprised all variants that had an LD r^2^ of ≥ 0.6 with an independent significant SNP. LD r^2^ values were obtained using the EUR 1000 Genomes Project Phase 3 reference panel. Gene-tissue expression analysis was performed with MAGMA v1.08 using GTEx v8 and BrainSpan tissue data (de Leeuw et al., 2015; The GTEx Consortium, 2020).

## Results

### Polygenic Overlap

Univariate MiXeR models indicated that there was sufficient power to perform bivariate MiXeR analyses, as all AIC values were positive. Full univariate results are provided in Supplementary Table 1. Bivariate model results are shown in Figure 1.

**Figure 1.**
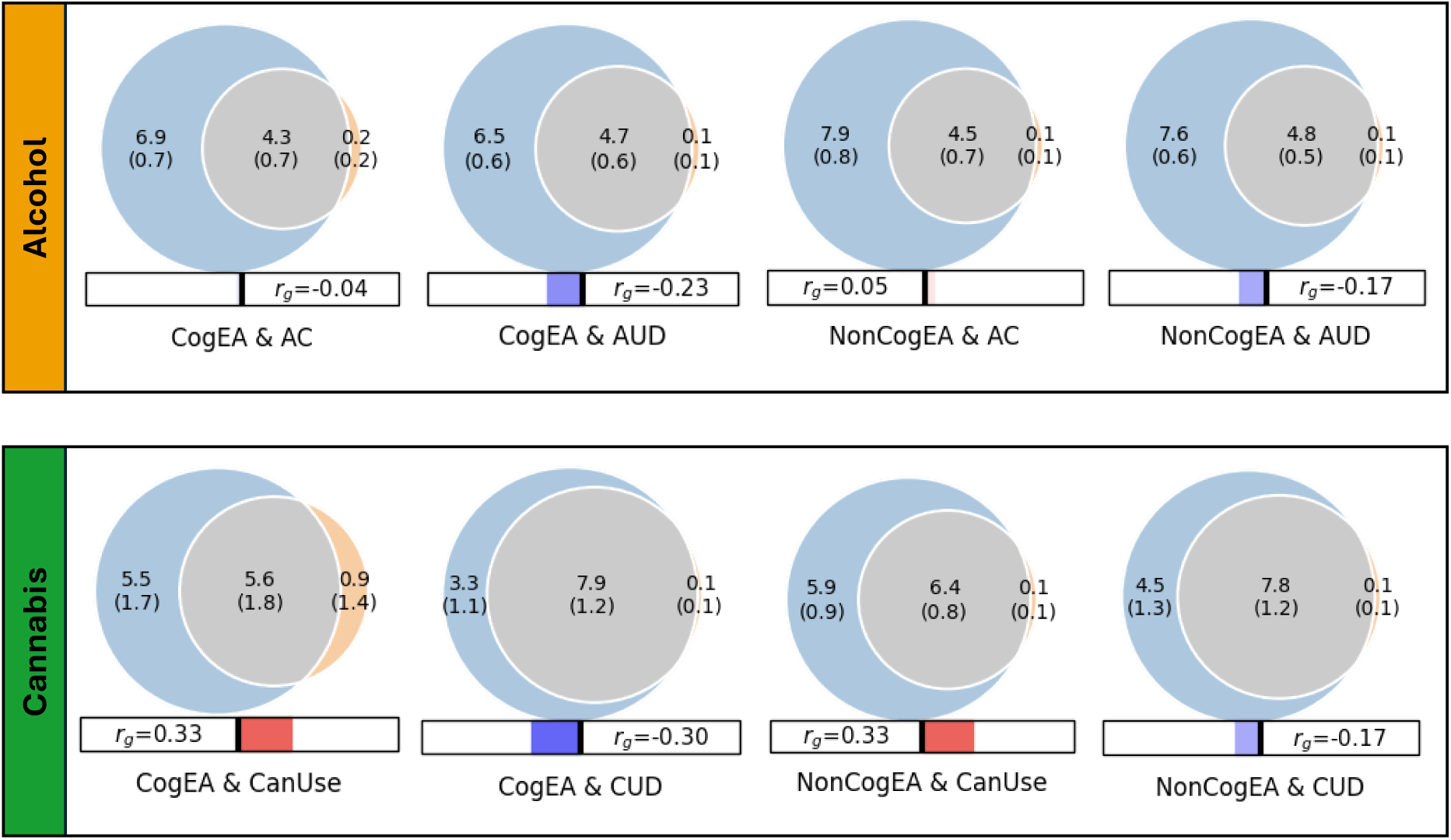
Results of bivariate causal mixture models (MiXeR). *Note:* CogEA = cognitive components of educational attainment, AC = alcohol consumption, AUD = alcohol use disorder, NonCogEA = non-cognitive components of educational attainment, CanUse = lifetime cannabis use, CUD = cannabis use disorder, r_g_ = genetic correlation.

#### Alcohol

EA shared a similar proportion of causal variants with AC (57.57%, SD = 0.07) and AUD (62.42%, SD = 0.04; Supplementary Figure 1). Despite similar polygenic overlap, over half of the shared variants for EA and AC had concordant effects (0.54, SD = 0.01), whereas concordance was lower for EA and AUD (0.37, SD = 0.01). CogEA and NonCogEA, which represent the cognitive and non-cognitive components of EA, respectively, showed moderate polygenic overlap with AC and AUD. Like with EA, however, effect directions differed. CogEA and AC shared 54.85% (SD = 0.06) of their causal variants, of which 48.12% (SD = 0.01) were concordant in direction, while CogEA and AUD shared 58.65% (SD = 0.05), with only 38.40% (SD = 0.01) concordant. NonCogEA showed a similar degree of overlap with both AC and AUD (52.55% and 55.23%, respectively), but more shared variants were directionally consistent with AC (52.86%, SD = 0.01) than AUD (41.02%, SD = 0.01). Notably, the vast majority of causal genetic variants for AC and AUD were shared by EA, CogEA, or NonCogEA.

#### Cannabis

In contrast to the alcohol-related traits, there was greater divergence in shared genetic effects of EA with CanUse and CUD. Although EA shared a lower proportion of causal variants with CanUse (48.07%, SD = 0.01) than CUD (84.18%, SD = 0.07; Supplementary Figure 2), the variants shared with CanUse were highly concordant (93.20%, SD = 0.03). Those shared with CUD, on the other hand, were much less concordant (38.30%, SD = 0.01). CogEA and NonCogEA exhibited a similar pattern. CogEA and CanUse shared 63.05% (SD = 0.18) of their causal variants, of which 71.42% (SD = 0.12) were directionally concordant. In contrast, for CogEA and CUD, although 81.66% of causal variants were shared, only 37.97% were concordant. Similarly, a smaller proportion of NonCogEA causal variants were shared with CanUse than with CUD (67.94%, SD = 0.06 vs. 76.67%, SD = 0.07), but a larger proportion of the variants shared with CanUse had concordant effect directions (65.56%, SD = 0.01 vs. 42.23%, SD = 0.01). Although CanUse exhibited more causal genetic variants that were not shared with EA (∼2.5k of 6.5k variants) than the other substance-related traits, most genetic variants for CanUse and CUD were shared with EA, CogEA, and NonCogEA.

### Local *r*_g_

#### Alcohol

Using LAVA, EA had significant local *r*_g_ with AC at 7 loci and with AUD at 6 loci, with one region (chr 14:99474534-100786189) overlapping between the two alcohol-related traits. Across all loci tested, most (66.86%) showed a concordant direction of *r*_g_ between EA and both AC and AUD. When examining EA subcomponents, CogEA showed more widespread local *r*_g_, including at 17 loci with AC and 23 loci with AUD. Five loci overlapped across the two alcohol-related traits and had the same direction of *r*_g_. Overall, 67.03% of all loci tested had a concordant direction of *r*_g_ across AC and AUD. NonCogEA showed fewer significant local *r*_g_ (Figure 2): 10 for AC and 3 for AUD, with no overlap. Nevertheless, most loci (63.32%) were concordant in direction of *r*_g_ across both alcohol traits. Full results are provided in Supplementary Tables 2-7.

**Figure 2.**
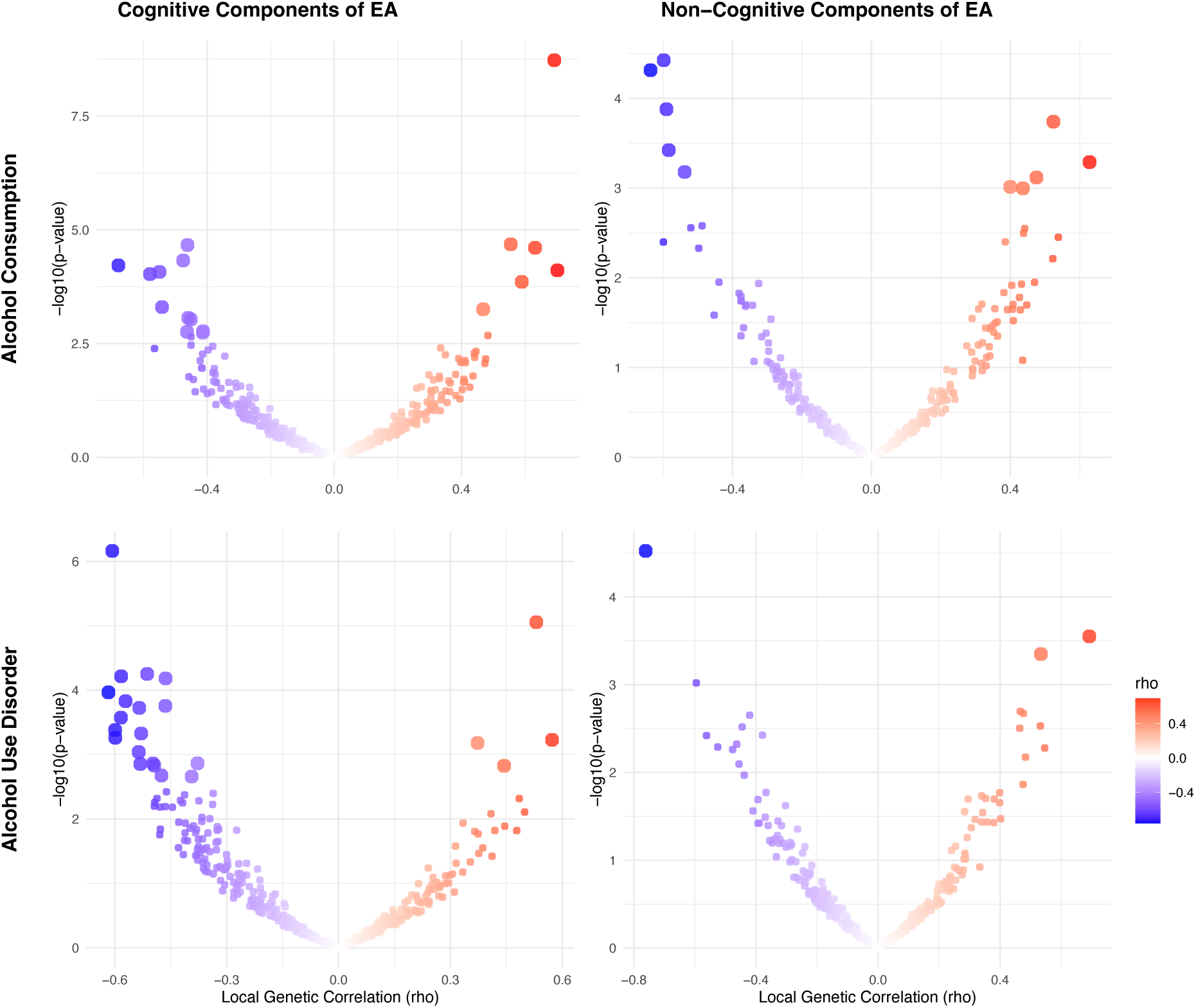
Volcano plots of local genetic correlations for alcohol consumption and use disorder. *Note:* EA = educational attainment. Larger dots represent significant local genetic correlations.

#### Cannabis

For cannabis traits, EA had significant *r*_g_ with CanUse at 17 loci and with CUD at 5 loci, though none of these loci were shared across the two cannabis-related traits. Just under half (44.64%) of all loci tested showed a concordant direction of *r*_g_ between EA and both CanUse and CUD, reflecting more divergence in local effects than for alcohol-related traits. For CogEA, there was significant local *r*_g_ with CanUse at 17 loci and with CUD at 7 loci, again with no overlap between the two. There were concordant directions of *r*_g_ for both cannabis traits with CogEA at 39.53% of loci. NonCogEA showed local *r*_g_ with CanUse at 9 loci and with CUD at 13, with no overlap. Concordance across the two cannabis-related traits with NonCogEA (42.59%) was slightly higher than for CogEA (39.53%). See Figure 3 for a summary of the results. Full results are in Supplementary Tables 8-13.

**Figure 3.**
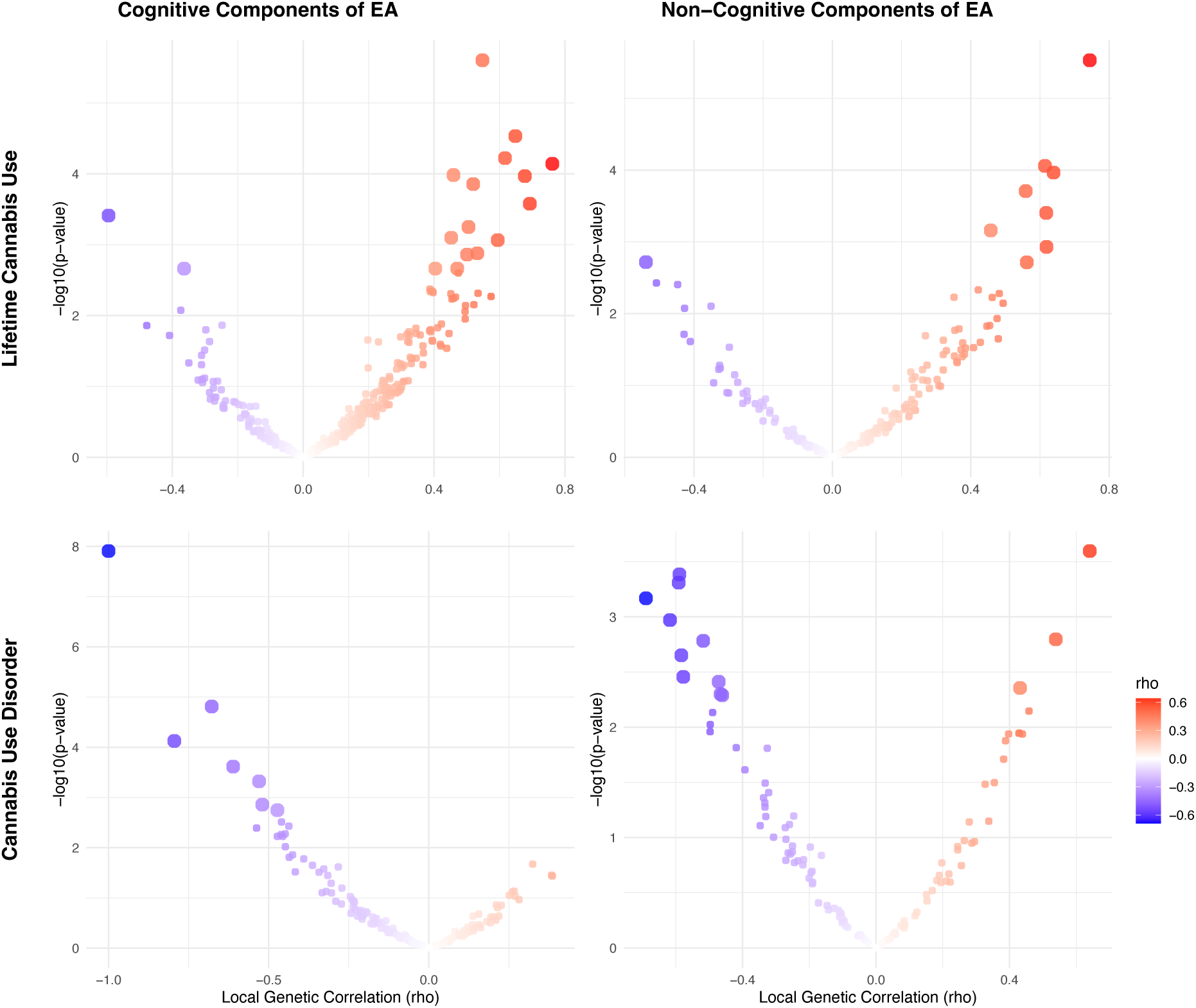
Volcano plots of local genetic correlations for lifetime cannabis use and disorder. *Note:* EA = educational attainment. Larger dots represent significant local genetic correlations.

### Joint SNP-Level Associations

#### Alcohol

ConjFDR analyses identified substantial overlap at the variant level between EA and alcohol-related traits. EA and AC had 108 jointly associated lead SNPs, of which 57 were concordant in effect direction. EA and AUD had 94 jointly associated SNPs, with 45 directionally concordant (Supplementary Figure 3). For CogEA, AC had 52 jointly associated lead SNPs (21 directionally concordant), while AUD had 67 (44 concordant). In contrast, NonCogEA had fewer joint associations with alcohol-related traits: 13 with AC (6 concordant) and 15 with AUD (6 concordant; Figure 4). Lead SNPs for the alcohol- and education-related trait pairs are in Supplementary Tables 14 and 15.

**Figure 4.**
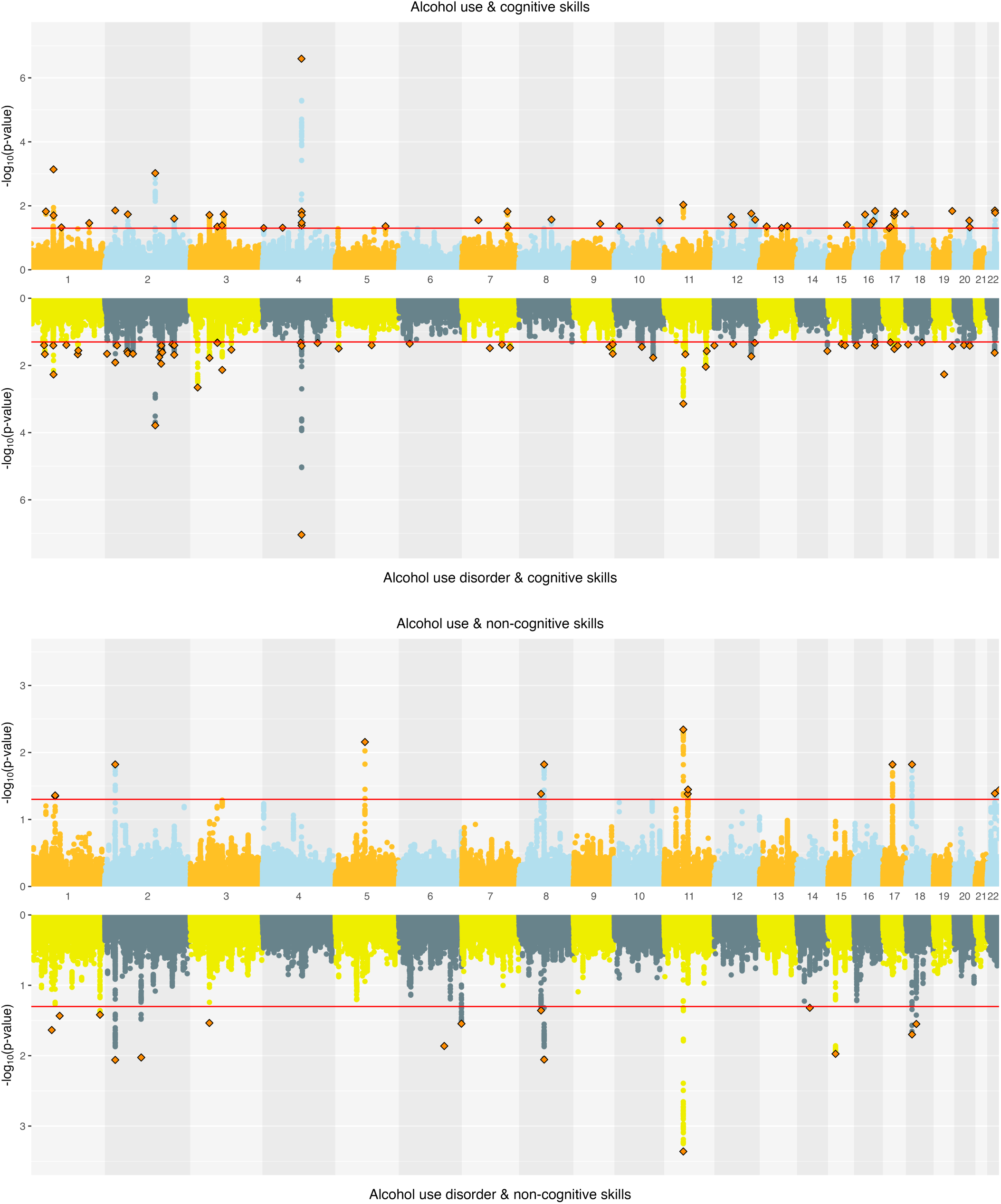
Miami plots of alcohol-related traits and educational attainment components.

#### Cannabis

Joint SNP-level associations were less abundant for cannabis-than alcohol-related traits but showed clearer separation between CanUse and CUD. EA and CanUse had 41 jointly associated SNPs, with 34 directionally concordant. EA and CUD had 28 jointly associated SNPs, with just 11 directionally concordant (Supplementary Figure 4). CogEA and CanUse had 36 jointly associated SNPs (13 concordant), while CogEA and CUD had 54 jointly associated SNPs (44 concordant), indicating greater overlap with CUD than CanUse. NonCogEA and CanUse had 11 jointly associated SNPs (4 concordant), while NonCogEA and CUD had 8 jointly associated SNPs (7 concordant; Figure 5), showing modest but highly directionally consistent associations of NonCogEA with CUD. Lead SNPs for the cannabis- and education-related trait pairs are in Supplementary Tables 16 and 17.

**Figure 5.**
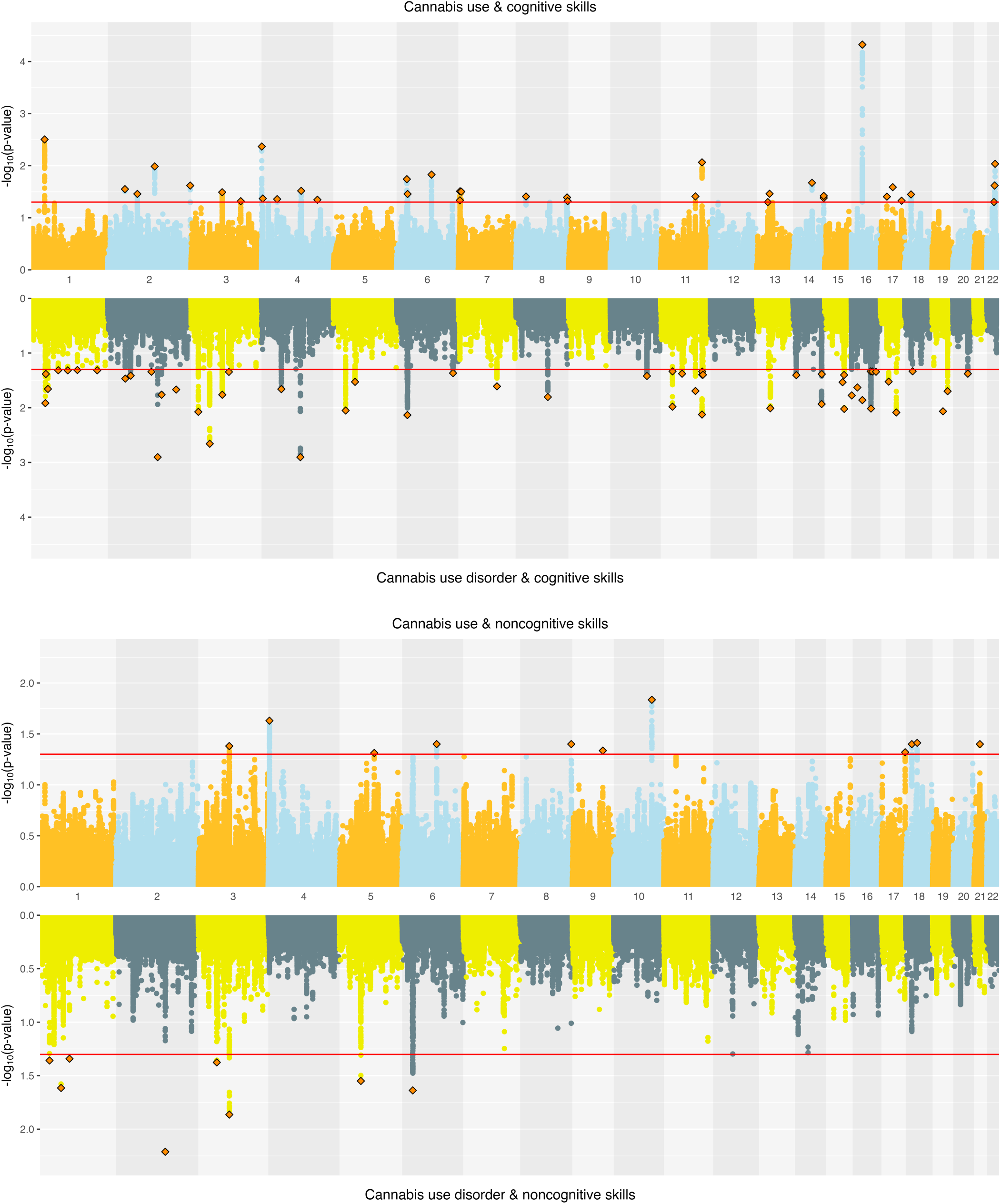
Miami plots of cannabis-related traits and educational attainment components.

### Functional Annotation

#### Alcohol

SNPs jointly associated with EA and both AC and AUD were enriched for expression in the brain during early, early-mid, and late-mid prenatal development. There was no significant enrichment for SNPs jointly associated with the alcohol traits and CogEA or NonCogEA. (Supplementary Figure 5). Analyses using MAGMA GTEx v8 tissues showed significant enrichment in brain and pituitary tissues for SNPs jointly associated with AUD and CogEA (Supplementary Figure 6), specifically in the nucleus accumbens (NAcc), hypothalamus, and caudate (Supplementary Figure 7).

#### Cannabis

For cannabis traits, only SNPs jointly associated with CanUse and EA showed significant enrichment during brain development in the early-mid and late-mid prenatal stages. No significant enrichment was identified for other jointly associated SNPs (Supplementary Figure 5). MAGMA tissue expression analysis showed significant SNP enrichment in skin tissues for CanUse and EA, in uterine cervix tissues for CanUse and CogEA, and in nerve tissues for CUD and CogEA (Supplementary Figure 6). The analysis across 54 tissues added specificity to the findings. For CanUse and EA, there was significant enrichment in the sun-exposed skin of the lower leg, and for CUD and CogEA, there was enrichment in the cervical spine and substantia nigra brain tissues. For CanUse and NonCogEA, there was enrichment in the cerebellum and its hemispheres (Supplementary Figure 7).

## Discussion

Across methods of increasing genetic specificity, our results provide insights into the paradox that EA is positively genetically correlated with use of alcohol or cannabis but negatively correlated with AUD and CUD. These directionally opposite *r*_g_ cannot be explained by differences in overall polygenic overlap as EA has greater polygenic overlap with AUD and CUD (62.42% and 84.18%) than with AC and CanUse (57.57% and 48.07%). However, the effect direction of the shared variants highlighted a key distinction. Variants shared between EA and SUDs more often had opposing effects on the two traits, while those shared with substance use had greater concordance.

Disaggregating EA into its cognitive (CogEA) and non-cognitive (NonCogEA) components clarified this distinction, with important nuance across methods. MiXeR estimated higher genome-wide concordance for CogEA with AC and CanUse than with AUD and CUD, consistent with the findings for EA overall. In contrast, conjFDR showed that the effect direction of jointly associated SNPs was more often concordant for CogEA and AUD/CUD than for CogEA and AC/CanUse. The discrepancy across the two methods is likely due to differences in scope. Whereas MiXeR estimates the polygenic overlap and concordance across *all* causal variants (including many with small effects), conjFDR identifies the SNPs that are most strongly jointly associated. Thus, although many variants positively associated with CogEA protect against developing AUD and CUD, a smaller set of variants with strong effects *increases* the risk for these disorders.

For NonCogEA, there were also differences across methods. MiXeR analyses showed higher concordance of NonCogEA with AC and CanUse than with AUD and CUD. For cannabis traits, however, conjFDR analysis showed the opposite pattern. SNPs jointly associated with NonCogEA and CUD were more directionally concordant than those jointly associated with NonCogEA and CanUse. Thus, although genetic variants associated with non-cognitive traits promote cannabis use generally, the most strongly associated variants may disproportionately contribute to risk for developing CUD. Variants may influence both NonCogEA (increasing liability for EA through non-cognitive traits) and CUD via risk-taking or socially adaptive forms of disinhibition (Moncel et al., 2025). In competitive academic environments, these tendencies facilitate goal pursuit and strategic risk-taking. For example, research shows that individuals in leadership positions often have elevated dominance (Lilienfeld et al., 2014; Lilienfeld et al., 2012) and that risk-taking is associated with better academic outcomes (Petzel & and Casad, 2022). However, in other contexts, these same traits can confer vulnerability to CUD.

Results from LAVA showed that genomic regions where EA has local *r*_g_ with AC/CanUse are largely distinct from those where EA is correlated with AUD/CUD. This pattern was consistent regardless of whether EA was considered as a whole or partitioned into cognitive and non-cognitive components. Previous research has also shown that experimentation with substances and the development of SUD are shaped by at least partially distinct etiological factors (Lynskey et al., 2003; Richmond-Rakerd et al., 2016). The minimal overlap identified in LAVA analyses suggests that transitions from use to disorder may involve a shift in biological pathways, such as from those associated with greater opportunity or openness to use substances to those linked with compulsivity and executive dysfunction, key elements of SUDs. Thus, local differences in pleiotropy may, in addition to differences in effect direction, contribute to the paradoxical associations of EA and its subcomponents with alcohol and cannabis use, and AUD and CUD.

SNP-level tissue enrichment analyses provided insight into the specific biological pathways that differentiate CanUse from CUD. Whereas SNPs jointly associated with CUD and CogEA were enriched in the substantia nigra, those jointly associated with CanUse and NonCogEA were enriched in the cerebellum and its hemispheres. Elevated dopamine function in the substantia nigra has been implicated in increased psychosis risk among individuals with CUD (Ahrens et al., 2025), and psychosis is often preceded by cognitive deficits (Jonas et al., 2022). These dopaminergic pathways in the substantia nigra may help explain the overlap between CUD and CogEA. In contrast, chronic cannabis use has been associated with altered cerebellar structure and poorer performance on non-cognitive behavioral tasks that involve the cerebellum (Blithikioti et al., 2019). Thus, the potentially distinct neurobiological pathways underlying substance use and SUDs may also differ in their associations with cognitive and noncognitive traits.

In contrast to the divergent effects between CanUse and CUD that were revealed by enrichment analyses, we found evidence of shared effects between AUD and AC. SNPs jointly associated with AUD and EA and those associated with AC and EA were linked to enriched expression in brain tissues during prenatal development. Prenatal stress plays a causal role in the development of SUDs later in life (Pastor et al., 2017) and is related to lower school achievement during childhood (Li et al., 2013). Our findings point to a shared genetic etiology that may account for this co-occurrence.

When EA was broken down into its subcomponents, SNPs jointly associated with AUD and CogEA showed enriched expression in the hypothalamus and two components of the basal ganglia—the NAcc and caudate nucleus. The basal ganglia have already been a target for intervention in AUD, with deep brain stimulation of the NAcc producing significant reductions in alcohol-related craving and compulsive behavior (Davidson et al., 2022). Other evidence supports the importance of these regions for understanding alcohol-related risk and cognitive functioning. For example, lower white matter integrity between the NAcc and prefrontal cortex is prospectively associated with earlier initiation of binge drinking (Morales et al., 2020). Similarly, the caudate nucleus is sensitive to prenatal alcohol exposure, with lower volume in this region predicting poorer cognitive control in individuals exposed to alcohol *in utero*, even after controlling for total brain volume, intelligence, and age (Fryer et al., 2012). Taken together, these findings suggest that genetic variants jointly associated with CogEA and AUD may influence risk by shaping the striatal circuits integral to balancing reward motivation with executive control.

Our study has several limitations. Analyses were conducted only among EUR individuals, which potentially limits generalizability to other genetically inferred ancestry groups. Educational systems, access to education, and the relationship between education and health differs across societies (Eikemo et al., 2008). Thus, the genetic architecture linking EA and substance-related traits may vary as well. Partitioning EA into CogEA and NonCogEA offered a more nuanced view of this complexity, but studies are needed to examine whether the observed patterns are consistent in other settings and groups. Additionally, although our analytic approach captured the overlap and divergence in genetic architecture for EA and substance-related traits, it does not provide insights into the causal directions of these associations.

By integrating multiple analyses of increasing genetic specificity, we show that the paradoxical genetic correlations between EA and alcohol/cannabis use versus AUD/CUD reflect more than just differences in overall polygenic overlap. Instead, a combination of differences in effect direction, associated genomic regions, and neurobiological pathways better explains the paradoxical associations. Partitioning EA into its cognitive and non-cognitive components highlights how traits generally viewed as promoting educational success—including curiosity, openness, and cognitive ability—may also increase vulnerability to the development of SUDs, particularly CUD. These findings reinforce our understanding of EA as a multi-dimensional trait with complex links to substance-related behaviors.

## Data Availability

All summary statistics used in the current study are publicly available. Summary statistics for alcohol consumption, alcohol use disorder, and cannabis use disorder can be accessed by applying for access to dbGaP accession phs001672. Lifetime cannabis use summary statistics were accessed via https://www.ru.nl/bsi/research/group-pages-0/substance-use-addiction-food-saf/vm-saf/genetics/international-cannabis-consortium-icc/. Educational attainment summary statistics can be accessed at: https://thessgac.com/. Summary statistics for the cognitive and non-cognitive components of educational attainment can be accessed at: NonCog GWAS: ftp://ftp.ebi.ac.uk/pub/databases/gwas/summary_statistics/GCST90011874, Cog GWAS: ftp://ftp.ebi.ac.uk/pub/databases/gwas/summary_statistics/GCST90011875.

## Acknowledgments

The views expressed in this article are those of the authors and do not necessarily represent the position or policy of the Department of Veterans Affairs, Uniformed Services University, the Department of Defense, or the US Government. This work was supported by the VISN 4 Mental Illness Research, Education, and Clinical Center.

## Disclosures

Dr. Kranzler is a member of advisory boards for Altimmune and Clearmind Medicine; a consultant to Sobrera Pharmaceuticals, Altimmune, and Lilly; the recipient of research funding and medication supplies for an investigator-initiated study from Alkermes and a company-initiated study by Altimmune; and an inventor on U.S. provisional patent “Multi-ancestry Genome-wide Association Meta-analysis of Buprenorphine Treatment Response.” The other authors have no disclosures to make.

