## Supplementary Materials for "Disaggregating the Genetic Overlap Between Educational Attainment and Substance Use Phenotypes"

### **Supplementary Figures**

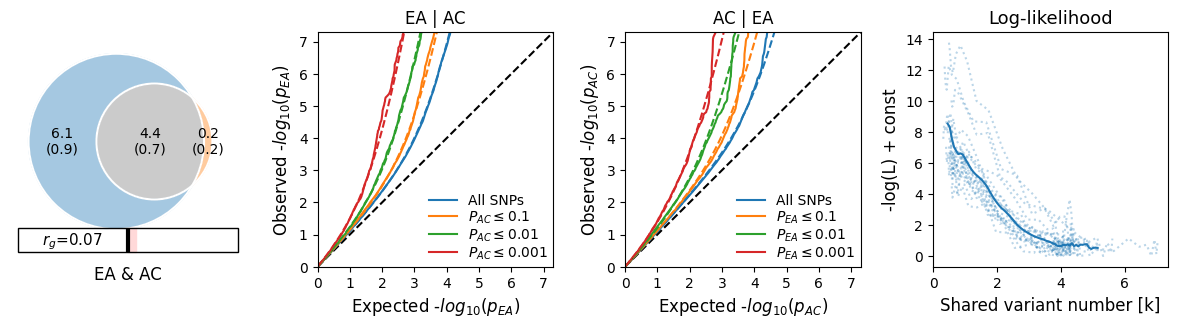

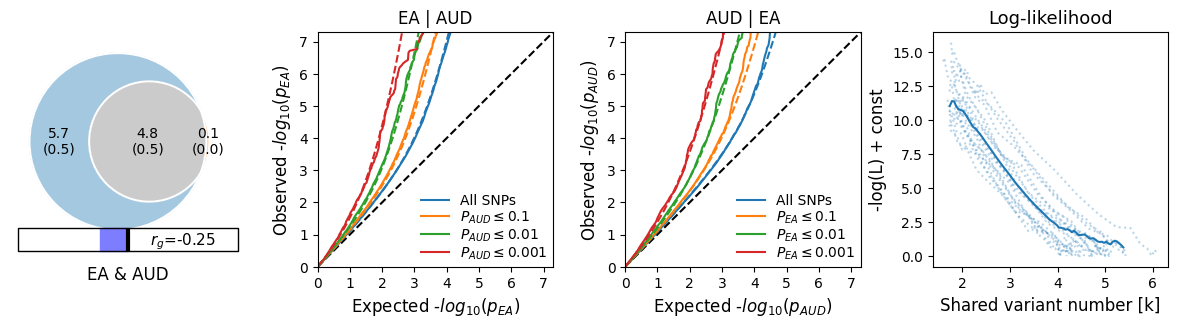

#### **Supplementary Figure 1. Bivariate MiXeR results for educational attainment (EA) and alcohol-related traits.**

*Note:* EA = educational attainment, AC = alcohol consumption, AUD = alcohol use disorder.

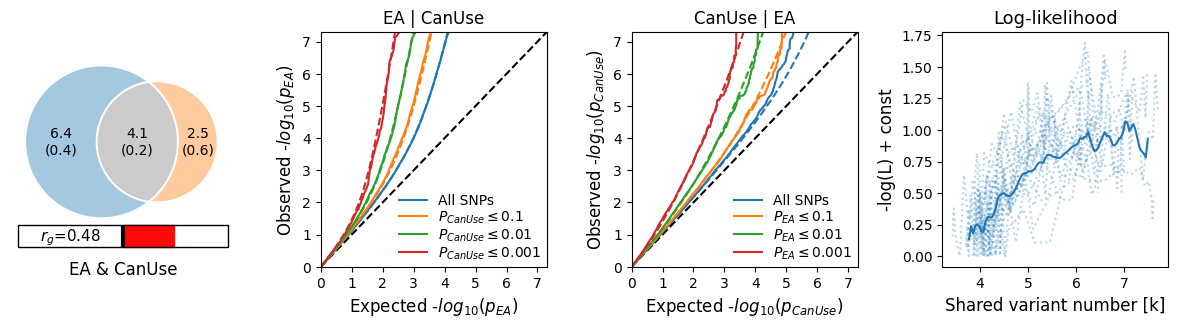

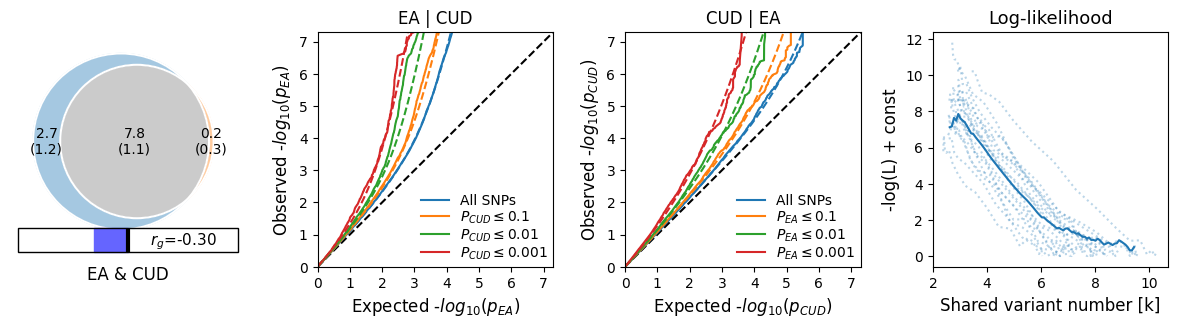

#### **Supplementary Figure 2. Bivariate MiXeR results for educational attainment (EA) and cannabis-related traits.**

*Note:* EA = educational attainment, CanUse = lifetime cannabis use, CUD = cannabis use disorder.

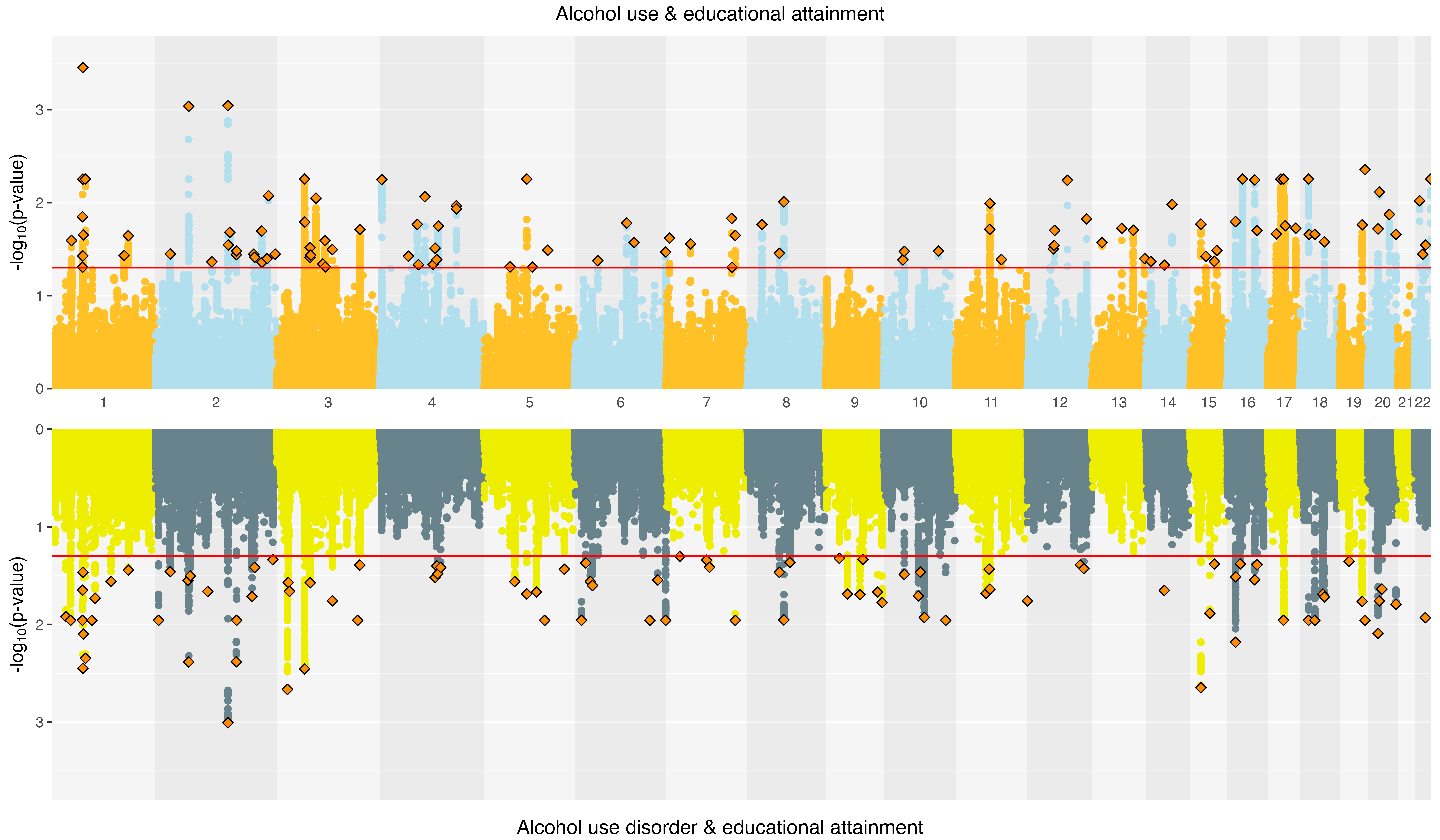

#### **Supplementary Figure 3. Miami plots of alcohol-related traits and educational attainment.**

*Note*: Log_10_-transformed conjFDR values for each SNP are on the y axes and chromosomal positions are on the x axes. The horizontal line is the threshold for significant shared associations of alcohol use and alcohol use disorder with educational attainment (conjFDR <0.05). Independent lead SNPs are outlined diamonds.

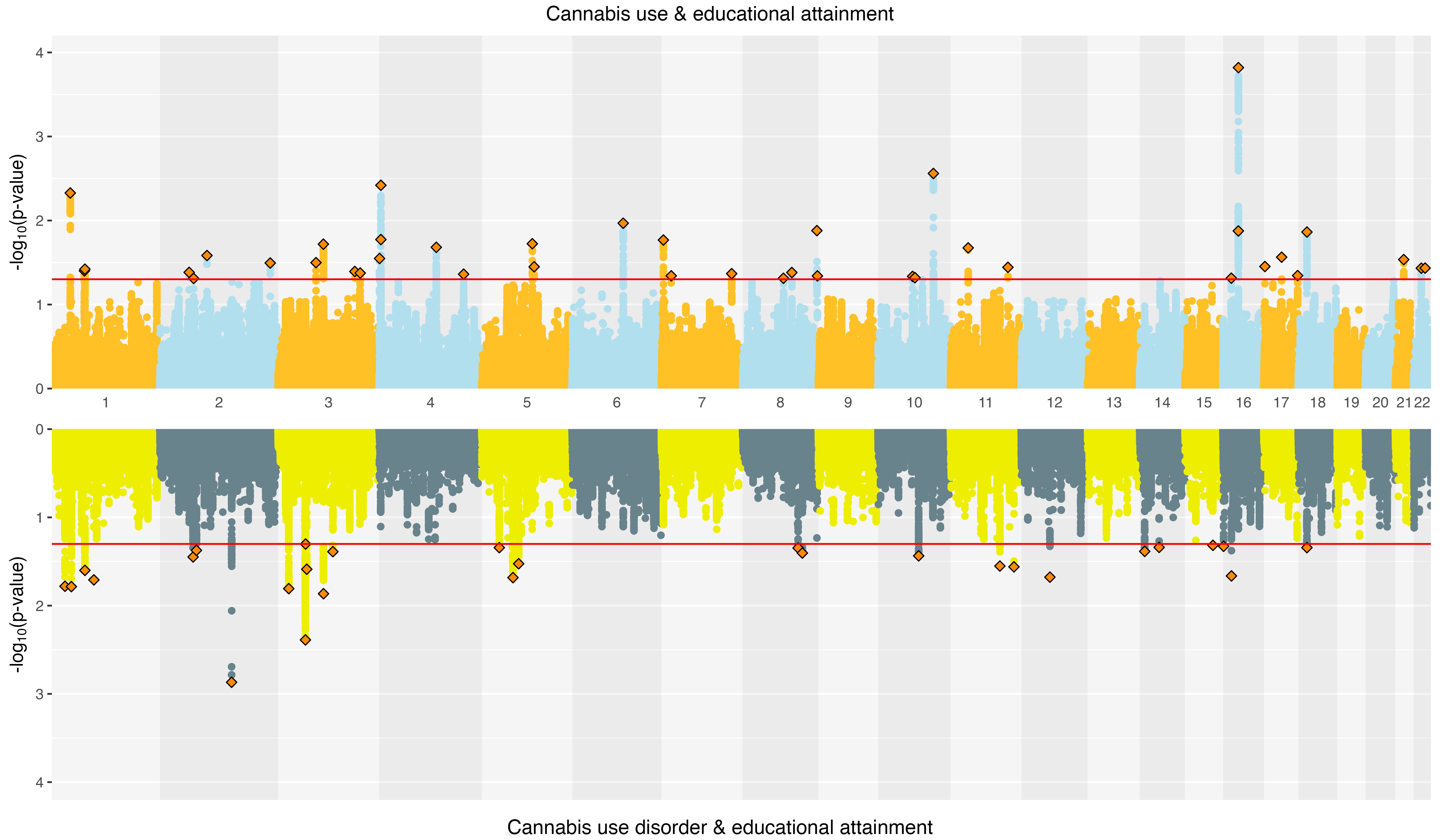

#### **Supplementary Figure 4. Miami plots of cannabis-related traits and educational attainment**

*Note*: Log_10_-transformed conjFDR values for each SNP are on the y axes and chromosomal positions are on the x axes. The horizontal line is the threshold for significant shared associations of cannabis use and cannabis use disorder with educational attainment (conjFDR <0.05). Independent lead SNPs are outlined diamonds.

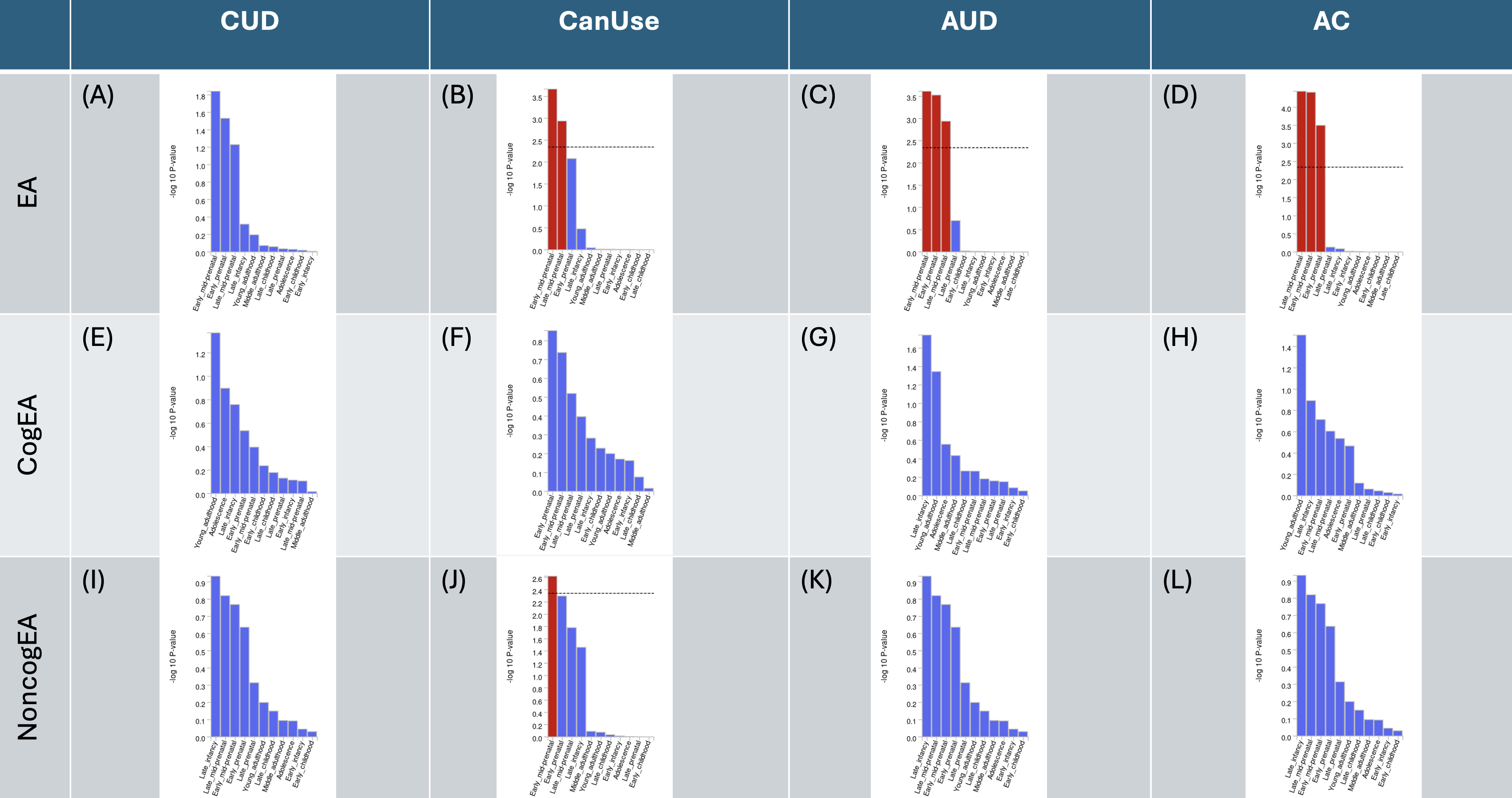

#### **Supplementary Figure 5. MAGMA results of BrainSpan 11 different developmental ages for each joint analysis.**

Results that were significant after multiple testing correction are highlighted in red. Panels A-D represent the results of enrichment analysis for joint condFDR analyses of EA with CUD, CanUse, AUD, and AC, respectively. E-H represent those of joint analyses of CogEA with the four substance-related traits, and I-L represent NonCogEA with the four traits.

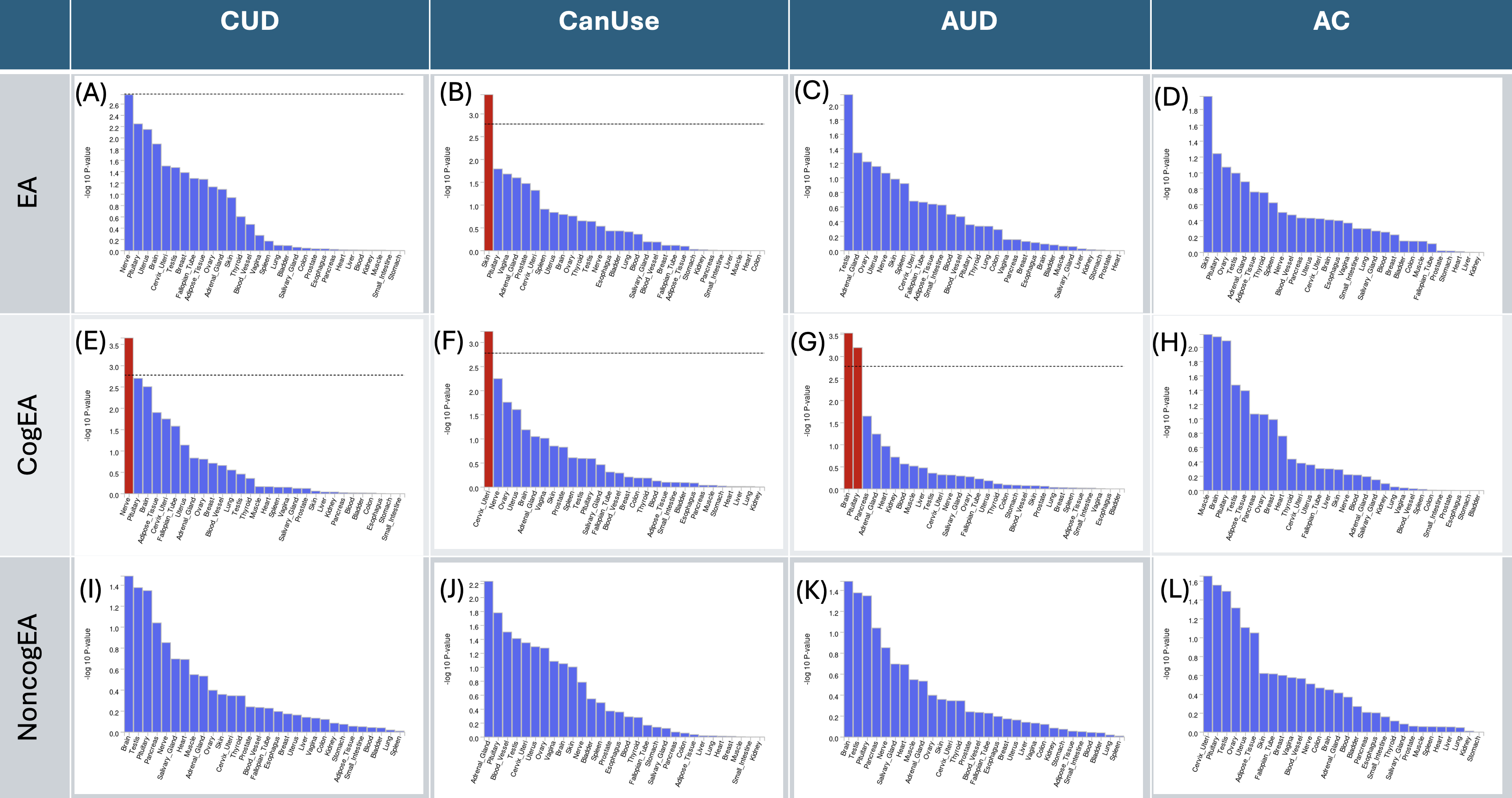

#### **Supplementary Figure 6. MAGMA results of GTEx v8 30 tissue types for each joint analysis.**

Results that were significant after multiple testing correction are highlighted in red. Panels A-D represent the results of enrichment analysis for joint condFDR analyses of EA with CUD, CanUse, AUD, and AC, respectively. E-H represent those of joint analyses of CogEA with the four substance-related traits, and I-L represent NonCogEA with the four traits.

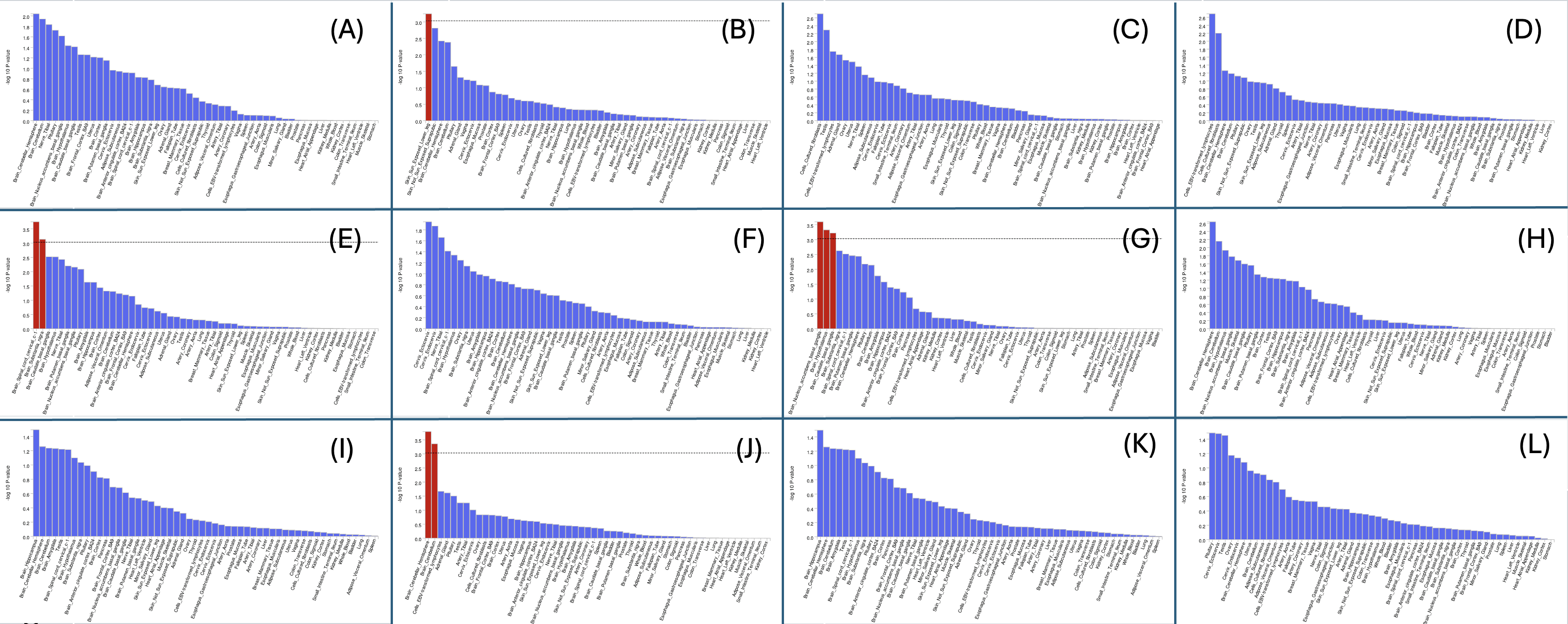

#### **Supplementary Figure 7. MAGMA results of GTEx v8 53 tissue types for each joint analysis.**

Results that were significant after multiple testing correction are highlighted in red. Panels A-D represent the results of enrichment analysis for joint condFDR analyses of EA with CUD, CanUse, AUD, and AC, respectively. E-H represent those of joint analyses of CogEA with the four substance-related traits, and I-L represent NonCogEA with the four traits.
